# Effect of peer-group participation on Knowledge about condoms among adolescent girls in rural Eastern Ethiopia: a community-based repeated cross-sectional study

**DOI:** 10.1101/2024.12.12.24318896

**Authors:** Nebiyou Fasil, Alemayehu Worku, Lemessa Oljira, Amare Worku Tadesse, Yemane Berhane

## Abstract

Adolescent girls are at high risk of acquiring sexually transmitted infections (STIs), including the human immunodeficiency virus (HIV). Global evidence indicates significance of comprehensive sexual education in empowering adolescents with accurate knowledge regarding safe sexual practices. Adolescents living in rural areas, particularly have inadequately developed life skills; limited health promotion knowledge and are at risk of being coerced into sexual relationships. The study aimed to assess the effect of peer group participation on improving knowledge about condoms among adolescent girls in rural eastern Ethiopia. The study analyzed data from a repeated cross-sectional study involving 3186 and 3290 adolescent girls during the baseline period (2016) and the endline period (2019), respectively. The respondents were adolescent girls aged 13–17 years old. The study’s outcome was knowledge about condoms, which was assessed using 3-item questions. A weighted multivariable logistic regression analysis was used to examine the effect of peer group participation on improving knowledge about condoms by controlling for potential confounders. Statistical significance was set at a p-value <0.05. The magnitude of good knowledge about condoms among peer-group participants was 23% higher in the endline compared to the baseline. The odds of having good knowledge about condoms among girls who participated in an organized peer group were 2.15 times higher than those who didn’t participate (AOR: 2.15, 95% CI: 1.42, 3.26). Moreover, the odds of having good knowledge about condoms among those who reported high confidence in perceived communication skills were 1.68 times higher than those who reported no or little confidence (AOR: 1.68, 95% CI: 1.33, 2.13). Peer-group participation improved knowledge about condoms. Girls with high confidence in their perceived communication skills also had better knowledge about condoms. Peer-group education interventions have the potential to improve condom knowledge and thus improve sexual and reproductive health outcomes of adolescent girls. Further studies are needed in other contexts to inform intervention scale-up.

## Introduction

Adolescence brings changes, including health challenges linked to the onset of sexual activity. Evidence related to adolescents’ physical health primarily focuses on adolescents over 15 years, while younger adolescents get little attention as they are considered too young (1). In sub-Saharan Africa, more girls than boys are sexually active at a younger age, within or outside the confines of marriage, with a corresponding increase in the risk of sexually transmitted infections (STIs), including Human Immunodeficiency Virus (HIV) (2–5). Condom use remains relatively low among adolescents despite extensive efforts (6).

Adolescent girls’ sexual and reproductive health (SRH) needs often go unmet due to lack of crucial health knowledge necessary for informed decision-making (7,8). Knowledge about condoms plays a critical role in promoting healthy sexual behavior by addressing misconceptions and misinformation that contribute to risky sexual behaviors (9).

Curriculum-based, theory-driven, group-based interventions targeting adolescent girls’ Knowledge and skills can influence their sexual behaviors (10–12). These interventions, grounded in behavioral frameworks are delivered through interactive group sessions enabling a structured and systematic delivery of information. Interventions addressing adolescents’ sexual and reproductive health outcomes involve individual, social, and environmental level interventions (13). Individual-level interventions primarily focus on knowledge and attitude to alter girls’ risk profiles (14,15). Peer group education, as part of multi-component interventions, yields mixed results (16–18), promoting a critical decision regarding the timing of these intervention.

Timing interventions during early adolescence or mid-late adolescence is crucial in devising appropriate strategies (19), with studies suggesting the effectiveness of early intervention (20). Sensitization on SRH issues peer education sessions have proven effective in preventing HIV and STIs (21,22).

Accurate Knowledge about condoms and HIV predicts preventive sexual behaviors (23). Conversely, misconceptions about condom use, including condom ineffectiveness, inaccessibility, and re-usability increase risks for pregnancy, STIs and other poor reproductive health outcomes (24).

Research on peer group interventions among very young adolescents in sub-Saharan Africa(25–27) is limited, reflecting the overall lack of attention to interventions in this demographic (28). Understanding the impact of interventions targeting adolescents is critical for meeting sustainable development goals(SDGs), Goal 3 on universal access to SRH services, including family planning, information and education to promote contraceptive use and prevent adolescent births (29); and African Union’s Agenda 2063, that seeks to address adolescent SRH needs (Somefun et al., 2021).

This study intends to uncover the factors that facilitate the acquisition of condom-related knowledge, providing vital insights for crafting targeted interventions aimed at improving sexual SRH outcomes among adolescent girls. With a burgeoning population of at-risk adolescents, it’s imperative to identify practical, adaptable, and cost-effective interventions catering to the SRH needs of adolescent girls, particularly in resource-constrained setting like Ethiopia. Inconsistent and non-continuous condom use persist in SSA countries, including Ethiopia. Thus, addressing knowledge-related issues stands as a crucial step in tackling sexual health risks, including unsafe sexual practices, a pressing public health concern for adolescents(31).

Therefore, this study aimed to examine the effect of peer-group participation on the improvement of knowledge about condoms among adolescent girls.

## Materials and Methods

### Study description

The study used data obtained from a quasi-experimental study called ‘Abdiboru’ project conducted as an implementation research project that assessed interventions to address structural issues facing adolescent girls in Western Hararghe zone, Oromia region, Eastern Ethiopia. The baseline survey was conducted from 31/05/2016 to 11/08/2016 and the endline survey was conducted from 09/10/2019 to 23/11/2019. The study population were adolescent girls aged 13-17 years old at the time of the surveys (Fig 1). Adolescent girls were sampled using a multi-stage sampling method, at first using simple random sampling method of clusters, followed by selection of households using computer generated random number that were identified after conducting complete census of the study area(32). For this study, 3186 and 3290 adolescent girls’ data were made available, from the baseline and endline surveys, respectively. The response rate for the baseline and endline surveys were 93.15% and 96.19%, respectively. Data for both surveys were collected using a structured and pretested interviewer-administered questionnaire that was translated into the local language, Affaan Oromo, for field use.

**Fig 1:**
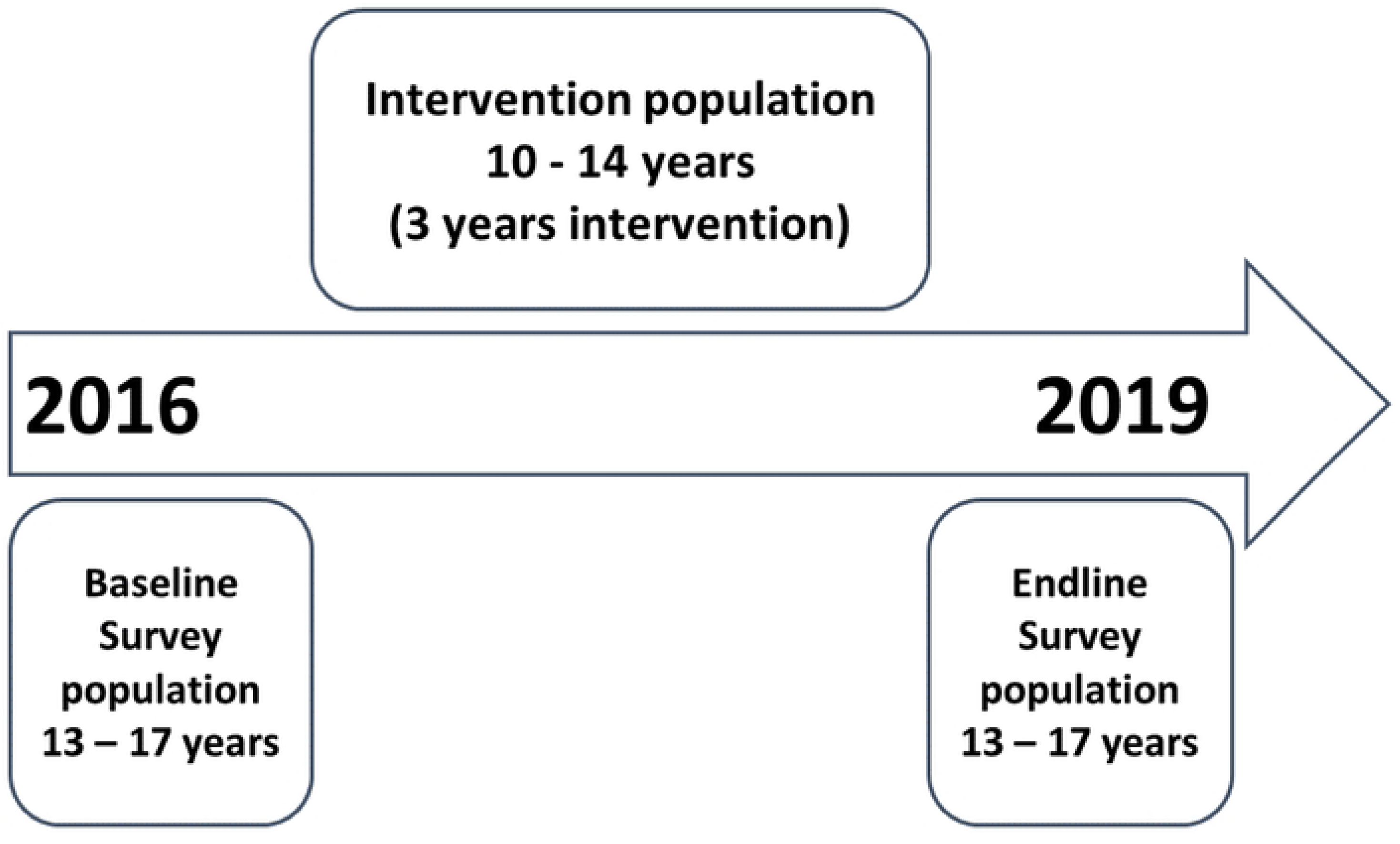
Intervention and survey populations and timing of surveys.

### About the peer-group participation intervention covered in the study

The peer groups were formed by 25-30 adolescent girls aged 10-14 years old who then went on to receive intervention for a three-year period (Figure 1). Each group upon formation, elected two representatives to facilitate their meetings. These two representatives participated in the training of trainers (TOTs) on the sexual and reproductive health (SRH) intervention curriculum prepared by the implementer, so they can go back to their groups and facilitate the discussions. Most peer groups meet every week, but some groups might also meet once every 2 weeks or once a month. Groups would meet in a permanent location, either in the school compound or in the village as per the groups’ consensus. The sexual and reproductive health intervention curriculum that was prepared and implemented had 11 discussion topics including, topics on Female Reproductive organ, Male reproductive organ, Puberty, Menstruation hygiene management, Pregnancy, Early marriage, Gender based violence, Family planning, STI (mode of transmission + prevention and treatment + HIV), Friendly health services, and the basic needs of adolescents. The groups would discuss each topic in each meeting session. Sessions were organized following the principles of participatory dialogue. They introduced materials and used participatory approaches such as role play, open-ended stories, and picture games to encourage dialogue on an equal footing within the group(33,34). The peer-group intervention was implemented in three districts, which served as the intervention arm. Another district that didn’t receive the implementation served as the control arm. After a three-year implementation period, the intervention population would grow to be 13-17 years old and serve as the study population for the endline survey.

### Measurement

#### Outcome variable

The outcome variable was Knowledge about condoms, assessed based on three questions. The three questions had “Yes”, “No” and “don’t know” response options. Adolescent girls who said “Yes” to questions “Condom can be used to prevent pregnancy” and “Condom can protect against getting HIV/AIDS and other sexually transmitted diseases; and also said “No” to the question “a condom can be reused” were considered to have good Knowledge about Condom.

#### Predictor variable

For exposure to peer-group participation intervention, an adolescent who lived in the intervention arm was considered as part of a peer group. Girls in the control arm were not considered part of the peer-group.

For peer group participation-Survey time interaction term, peer group participation(P1) was labeled as “1” and not participating(P0) was labeled as “0”. With regards to survey time, baseline(T0) was labelled as “0” and endline(T1) was labeled as “1”. The interaction term was constructed by combining these variables; hence adolescents who lived in the intervention arm and were assessed during the endline time(P1T1) were labeled as “1(Yes)”, Girls who lived in the control arm and were assessed during the baseline time(P0T0) and endline time (P0T1), and intervention arm participants assessed during the baseline (P1T0) were labeled as “0(No)”.

#### Covariates

Perceived communication skill level was assessed using a single-item question about their perceived level of confidence and adolescent girls responded as “no/little confidence”, “moderate confidence” and “high confidence”(35).

Health Extension workers(HEW) are community health workers who are tasked with providing basic curative, promotive and preventive services by conducting home and school visits(36). Reported ever having had contact with HEW was considered as one of the covariates.

### Data analysis

Data were analyzed using STATA/SE 14.0. Descriptive analysis was conducted to assess the magnitude of knowledge about condoms as per adolescent girl’s peer group participation and socio-demographic and other behavioral characteristics. Multivariable regression analysis controlled for covariates such as Adolescent girls’ age, education, marital status and contact with health extension workers. Intervention, survey time, intervention-survey time interaction term was also included in the analysis to estimate the effect of the intervention on the outcome. Intervention effect was examined using Odds ratio with 95% confidence intervals, and statistical significance was declared at p<0.05. Appropriate weighting was done to account for the complex survey design and analysis.

### Ethics approval

The original study protocol was approved by the Institutional Review Board of Addis Continental Institute of Public Health (Ref No. ACIPH/IRB/005/2016). Informed verbal consent was obtained from all study participants in the presence of parents/guardians. For participants below the age of 15 years, additional parental/guardian-informed verbal consent was obtained. All interviews took place in a private setting to assure confidentiality. Permission for data use was sought and secured from the main study principal investigator. No personal identifiers were linked with the dataset that was made available and utilized for this study.

## Results

Most respondents in both surveys attended primary school (49.2% and 54.7%) and were not married (88.01% and 93.25%). (Table 1)

**Table 1:** Description of the study population (Weighted)

Nearly eight out of ten (76.83%) adolescents reported moderate to high confidence in their communication during the baseline, while almost nine out of ten girls reported the same in the endline. In both surveys, only one out of ten girls reported ever having contact with health extension workers (Community health workers) (Table 2).

**Table 2:** Perceived communication skill and contact with HEW characteristics of adolescent girls of West Hararghe, 2016 and 2019 (Weighted)

In terms of intervention status, the magnitude of good Knowledge among peer-group participants during endline was 23 percent higher compared to the baseline, while among the non-peer-group, there was a decrease. (Table 3)

**Table 3:** Good Condom knowledge and peer group participation of adolescent girls (Weighted percentage)

The weighted multivariable logistic regression analysis showed that the odds of having good Knowledge about condoms among girls who had peer-group participation assessed during the endline time was 2.15 times higher than those who didn’t participate and were assessed during the baseline time (Adjusted OR: 2.156, 95% CI: 1.425, 3.26). Moreover, the odds of having good Knowledge about condoms among those who reported high confidence in perceived communication skills were 1.68 times higher than those who reported no/little confidence (Adjusted OR: 1.682, 95% 1.329, 2.129). (Table 4)

**Table 4:** Multivariable logistic regression analysis on the effect of peer-group participation intervention on Knowledge about Condom (Weighted)

## Discussion

The objective of this study was to assess how peer-group involvement impacts the enhancement of condom-related knowledge among adolescent girls. Peer-group participation significantly improved knowledge level on condoms among adolescents. Knowledge about condoms was also higher among girls who reported moderate to high confidence in their communication skill. Adolescent girls who had peer-group education had better Knowledge about condoms. This finding aligns with established literature demonstrating the effectiveness of peer-based education in improving HIV risk reduction knowledge, particularly regarding condom use (37). Studies conducted in India(38,39), Nigeria(26), and Ghana (40) further corroborate this result. Notably, systematic reviews from Western Pacific region also highlight the significant enhancement of adolescents’ knowledge through peer education (41).

Peer-group interventions offer a conductive platform for discussing sensitive issues, facilitating more comfortable communication among adolescent girls compared to adult-led interventions(16). Researches underscore the effectiveness of intervening during early adolescence in low and middle-income countries, highlighting the improvement in health mediators and health behaviors(42,43). Such forums enable knowledge sharing among adolescent girls, which has been effective in sub-Saharan countries, especially within school-based peer-group interventions (23,27,44,45). Moreover, establishing Girls’ clubs locally can serve as an entry point for the implementing impactful interventions without being resource intensive. Initial work is important in training students, to lead discussions within these clubs, coupled with teachers’ oversight and support, presents a feasible and sustainable approach.

However, the plight of out-of-school adolescent girls remains overlooked, with limited evidence of community-based interventions addressing their needs (17,26). Lack of access to comprehensive sexual education, limited access to health services and economic vulnerabilities are risks that make these girls more vulnerable. Addressing adolescent girls in these low-resource settings necessitates resource allocation in identifying, organizing, training, and monitoring activities, typically by diverse bodies such as NGOs community, or local organizations. Furthermore, creating cost-effective and sustainable interventions becomes pivotal in such contexts, often requiring community centers as meeting points for adolescent girls.

Furthermore, the study emphasizes the importance of communication skills in influencing adolescent girls’ knowledge about condoms. Studies from South Africa and Colombia underscored the pivotal role of good communication skills in fostering protective behavior(23) and addressing gaps in sexual health knowledge(46).

Notably, the study’s strength lies in its employment of a significant sample size, ensuring robustness in evaluating intervention impacts. Additionally, the research delved into early intervention effects on adolescent sexual health knowledge, an area often overlooked. However, the study’s limitation is its inability to assess condom use timing and correct application, components of knowledge that could have enriched the depth of knowledge assessment and produced a comprehensive evaluation of knowledge depth.

## Conclusion

In conclusion, peer-group participation improved adolescent girls’ knowledge about condoms. In addition, condom-related knowledge was associated with good perceived communication skills. Peer-group education interventions have the potential to improve condom knowledge and thus improve sexual and reproductive health outcomes of adolescent girls. Further studies are needed to evaluate the effectiveness of the intervention in other contexts to inform intervention scale-up and adoption into national adolescent health promotion strategies.

## Data Availability

All data relevant to the study are included in the manuscript. The data can also be accessible upon request.

## Acknowledgements

The authors would like to acknowledge the Abdiboru project for providing the data for this paper.

## References

1. Jones N, Pincock K, Baird S, Yadete W, Hamory Hicks J. Intersecting inequalities, gender and adolescent health in Ethiopia. Int J Equity Health. 2020 Jun 15;19:97.

2. Dlamini BR, Mabuza P, Thwala Tembe M, Masangane Z, Dlamini P, Simelane E. Are Adolescents and Youth Programs Missing The Real Targets? Analysis of Socio-Cultural Factors Influencing Use of Sexual Reproductive Health Services by Young People in Swaziland. J AIDS Clin Res [Internet]. 2017 [cited 2019 Aug 28];08(04). Available from: https://www.omicsonline.org/open-access/are-adolescents-and-youth-programs-missing-the-real-targets-analysis-of-sociocultural-factors-influencing-use-of-sexual-reproducti-2155-6113-1000684.php?aid=88254

3. Millanzi WC, Kibusi SM, Osaki KM. Effect of integrated reproductive health lesson materials in a problem-based pedagogy on soft skills for safe sexual behaviour among adolescents: A school-based randomized controlled trial in Tanzania. PLoS One. 2022 Feb 22;17(2):e0263431.

4. Mmbaga EJ, Kajula L, Aarø LE, Kilonzo M, Wubs AG, Eggers SM, et al. Effect of the PREPARE intervention on sexual initiation and condom use among adolescents aged 12–14: a cluster randomised controlled trial in Dar es Salaam, Tanzania. BMC Public Health. 2017 Apr 17;17:322.

5. Chapman J, do Nascimento N, Mandal M. Role of Male Sex Partners in HIV Risk of Adolescent Girls and Young Women in Mozambique. Glob Health Sci Pract. 2019 Sep 23;7(3):435–46.

6. Bankole A, Ahmed FH, Neema S, Ouedraogo C, Konyani S. Knowledge of correct condom use and consistency of use among adolescents in four countries in Sub-Saharan Africa. Afr J Reprod Health. 2007;11(3):197–220.

7. Salam RA, Faqqah A, Sajjad N, Lassi ZS, Das JK, Kaufman M, et al. Improving Adolescent Sexual and Reproductive Health: A Systematic Review of Potential Interventions. J Adolesc Health. 2016 Oct;59(4 Suppl):S11–28.

8. Jisso M, Feyasa MB, Medhin G, Dadi TL, Simachew Y, Denberu B, et al. Sexual and reproductive health service utilization of young girls in rural Ethiopia: What are the roles of health extension workers? Community-based cross-sectional study. BMJ Open. 2022 Sep 21;12(9):e056639.

9. Kanda L, Mash R. Reasons for inconsistent condom use by young adults in Mahalapye, Botswana. Afr J Prim Health Care Fam Med. 2018 May 24;10(1):1492.

10. Rosenberg NE, Gichane MW, Vansia D, Phanga T, Bhushan NL, Bekker LG, et al. Assessing the Impact of a Small-Group Behavioral Intervention on Sexual Behaviors Among Adolescent Girls and Young Women in Lilongwe Malawi: A Quasi-Experimental Cohort Study. AIDS Behav. 2020 May 1;24(5):1542–50.

11. Anna Downie. Our Theory of change: For sustaining community action on HIV, health and rights [Internet]. International HIV/AIDS Alliance; 2017. Available from: https://reliefweb.int/sites/reliefweb.int/files/resources/alliance_toc-2017_original.pdf

12. Plourde KF, Ippoliti NB, Nanda G, McCarraher DR. Mentoring Interventions and the Impact of Protective Assets on the Reproductive Health of Adolescent Girls and Young Women. Journal of Adolescent Health. 2017;30:1e9.

13. Visser M. Rethinking HIV-prevention for school-going young people based on current behaviour patterns. SAHARA J. 2017 Sep 21;14(1):64–76.

14. Wamoyi J, Mshana G, Mongi A, Neke N, Kapiga S, Changalucha J. A review of interventions addressing structural drivers of adolescents’ sexual and reproductive health vulnerability in sub-Saharan Africa: implications for sexual health programming. 2014;

15. Sommer M, Mmari K. Addressing Structural and Environmental Factors for Adolescent Sexual and Reproductive Health in Low- and Middle-Income Countries. Am J Public Health. 2015 Oct;105(10):1973–81.

16. Siddiqui M, Kataria I, Watson K, Chandra-Mouli V. A systematic review of the evidence on peer education programmes for promoting the sexual and reproductive health of young people in India. Sex Reprod Health Matters. 2020;28(1):1741494.

17. Rose-Clarke K, Bentley A, Marston C, Prost A. Peer-facilitated community-based interventions for adolescent health in low- and middle-income countries: A systematic review. PLoS One. 2019 Jan 23;14(1):e0210468.

18. Dodd S, Widnall E, Russell AE, Curtin EL, Simmonds R, Limmer M, et al. School-based peer education interventions to improve health: a global systematic review of effectiveness. BMC Public Health. 2022 Dec 2;22(1):2247.

19. Stanton B, Dinaj-Koci V, Wang B, Deveaux L, Lunn S, Li X, et al. Adolescent HIV risk reduction in The Bahamas: Results from two randomized controlled intervention trials spanning elementary school through high school. AIDS Behav. 2016 Jun;20(6):1182–96.

20. Maticka-Tyndale E, Wildish J, Gichuru M. Quasi-experimental evaluation of a national primary school HIV intervention in Kenya. Evaluation and Program Planning. 2007;30(2):172.

21. Jennings L, George AS, Jacobs T, Blanchet K, Singh NS. A forgotten group during humanitarian crises: a systematic review of sexual and reproductive health interventions for young people including adolescents in humanitarian settings. Confl Health. 2019 Nov 27;13:57.

22. He J, Wang Y, Du Z, Liao J, He N, Hao Y. Peer education for HIV prevention among high-risk groups: a systematic review and meta-analysis. BMC Infect Dis. 2020 May 12;20:338.

23. Harrison A, Smit J, Hoffman S, Nzama T, Leu CS, Mantell J, et al. Gender, peer and partner influences on adolescent HIV risk in rural South Africa. Sex Health. 2012 May;9(2):178–86.

24. Mbachu CO, Agu IC, Obayi C, Eze I, Ezumah N, Onwujekwe O. Beliefs and misconceptions about contraception and condom use among adolescents in south-east Nigeria. Reprod Health. 2021 Jan 6;18:7.

25. Manda WC, Pilgrim N, Kamndaya M, Mathur S, Sikweyiya Y. Girl-only clubs’ influence on SRH knowledge, HIV risk reduction, and negative SRH outcomes among very young adolescent girls in rural Malawi. BMC Public Health. 2021 Apr 27;21:806.

26. Akuiyibo S, Anyanti J, Idogho O, Piot S, Amoo B, Nwankwo N, et al. Impact of peer education on sexual health knowledge among adolescents and young persons in two North Western states of Nigeria. Reprod Health. 2021 Oct 12;18:204.

27. Ross DA, Changalucha J, Obasi AI, Todd J, Plummer ML, Cleophas-Mazige B, et al. Biological and behavioural impact of an adolescent sexual health intervention in Tanzania: a community-randomized trial. AIDS. 2007 Sep 12;21(14):1943–55.

28. Habte A, Dessu S, Bogale B, Lemma L. Disparities in sexual and reproductive health services utilization among urban and rural adolescents in southern Ethiopia, 2020: a comparative cross-sectional study. BMC Public Health. 2022 Jan 31;22:203.

29. Mason-Jones AJ, Sinclair D, Mathews C, Kagee A, Hillman A, Lombard C. School-based interventions for preventing HIV, sexually transmitted infections, and pregnancy in adolescents. Cochrane Database Syst Rev. 2016 Nov 8;2016(11):CD006417.

30. Somefun OD, Casale M, Haupt Ronnie G, Desmond C, Cluver L, Sherr L. Decade of research into the acceptability of interventions aimed at improving adolescent and youth health and social outcomes in Africa: a systematic review and evidence map. BMJ Open. 2021 Dec 20;11(12):e055160.

31. Yosef T, Nigussie T. Behavioral Profiles and Attitude toward Condom Use among College Students in Southwest Ethiopia. Biomed Res Int. 2020;2020:9582139.

32. Berhane Y, Worku A, Tewahido D, Fasil N, Gulema H, Tadesse AW, et al. Adolescent Girls’ Agency Significantly Correlates With Favorable Social Norms in Ethiopia—Implications for Improving Sexual and Reproductive Health of Young Adolescents. J Adolesc Health. 2019 Apr;64(4 Suppl):S52–9.

33. ACIPH, CARE Ethiopia, Bill and Melinda Gates Foundation. Abdiboru Project Improving Adolescent Reproductive Health and Nutrition through Structural Solutions: Midterm Report [Internet]. 2018 Nov [cited 2022 Dec 4] p. 43. Available from: https://www.careevaluations.org/evaluation/abdiboru-project-improving-adolescent-reproductive-health-and-nutrition-through-structural-solutions-midterm-report/

34. Edmeades J, Lantos H, Mekuria F. Worth the effort? Combining sexual and reproductive health and economic empowerment programming for married adolescent girls in Amhara, Ethiopia. 2016;

35. Fasil N, Worku A, Oljira L, Tadesse AW, Berhane Y. Association between sexual and reproductive health education in peer group and comprehensive knowledge of HIV among adolescent girls in rural eastern Ethiopia: a community-based cross-sectional study. BMJ Open. 2022 Oct 1;12(10):e063292.

36. Assefa Y, Gelaw YA, Hill PS, Taye BW, Van Damme W. Community health extension program of Ethiopia, 2003–2018: successes and challenges toward universal coverage for primary healthcare services. Globalization and Health. 2019 Mar 26;15(1):24.

37. Faust L, Yaya S. The effect of HIV educational interventions on HIV-related knowledge, condom use, and HIV incidence in sub-Saharan Africa: a systematic review and meta-analysis. BMC Public Health. 2018 Nov 13;18(1):1254.

38. Parwej S, Kumar R, Walia I, Aggarwal AK. Reproductive health education intervention trial. Indian J Pediatr. 2005 Apr;72(4):287–91.

39. Barua A, Watson K, Plesons M, Chandra-Mouli V, Sharma K. Adolescent health programming in India: a rapid review. Reproductive Health. 2020 Jun 3;17(1):87.

40. Geugten J van der, Meijel B van, Uyl MHG den, Vries NK de. Evaluation of a Sexual and Reproductive Health Education Programme: Students’ Knowledge, Attitude and Behaviour in Bolgatanga Municipality, Northern Ghana. African Journal of Reproductive Health. 2015 Oct 29;19(3):126–36.

41. Xu T, Tomokawa S, Gregorio ER, Mannava P, Nagai M, Sobel H. School-based interventions to promote adolescent health: A systematic review in low- and middle-income countries of WHO Western Pacific Region. PLoS One. 2020 Mar 5;15(3):e0230046.

42. Hallman K, Stoner M, Chau M, Melnikas AJ. A review of control-comparison interventions on girls and health in low and middle-income countries. Girl Hub. 2013;

43. Austrian K, Soler-Hampejsek E, Kangwana B, Wado YD, Abuya B, Maluccio JA. Impacts of two-year multisectoral cash plus programs on young adolescent girls’ education, health and economic outcomes: Adolescent Girls Initiative-Kenya (AGI-K) randomized trial. BMC Public Health. 2021 Nov 24;21:2159.

44. Barnett S, Dijk J van, Swaray A, Amara T, Young P. Redesigning an education project for child friendly radio: a multisectoral collaboration to promote children’s health, education, and human rights after a humanitarian crisis in Sierra Leone. BMJ. 2018 Dec 7;363:k4667.

45. Lameiras-Fernández M, Martínez-Román R, Carrera-Fernández MV, Rodríguez-Castro Y. Sex Education in the Spotlight: What Is Working? Systematic Review. Int J Environ Res Public Health. 2021 Mar 4;18(5):2555.

46. Morales A, Vallejo-Medina P, Abello-Luque D, Saavedra-Roa A, García-Roncallo P, Gomez-Lugo M, et al. Sexual risk among Colombian adolescents: knowledge, attitudes, normative beliefs, perceived control, intention, and sexual behavior. BMC Public Health. 2018 Dec 17;18:1377.

